# Computerized biofeedback to characterize Pupil Cycle Time (PCT) in neuropathies and retinopathies

**DOI:** 10.1101/2022.05.24.22275446

**Authors:** Suzon Ajasse, Catherine Vignal-Clermont, Saddek Mohand-Saïd, Cecilia Coen, Carole Romand, Jean Lorenceau

## Abstract

Pupillary responses to light offer a convenient and objective way to quickly assess the functional health of the anterior afferent visual pathways. We here present the characteristics of Pupil Cycle Time (PCT) obtained with a computerized biofeedback setting in patients with retinal and optic nerve diseases. The spectral analysis of the sustained pupillary oscillations elicited over 45 seconds of passive fixation of colored displays with different spatial configurations provides relevant information that allow distinguishing patients from healthy participants with good sensitivity and specificity. PCT measures done with this method could complement the current functional examination of chronic ophthalmic diseases whose prevalence worryingly increases worldwide.

## Introduction

The functional evaluation of vision is an important step in diagnosing and following-up chronic ophthalmic diseases whose prevalence worryingly increases worldwide^1,2^. As a general rule, individuals consult only after they felt a visual loss, which often occurs after structural damages developed^3^. The functional evaluation of vision is performed using subjective tests where individuals are asked to report on their visual percepts (for instance to assess visual acuity, or to perform Standard Automated Perimetry, SAP). Both the patients and the practitioners agree to consider that SAP is tedious and long^4^. In addition, the results can be hampered by fluctuations of attention, eye-movements, fatigue, or stress. As a consequence SAP is variable^5^, limiting its usefulness for following-up the evolution of a disease. Attempts to shorten the SAP exam have been made^6^, but these do not avoid some of the above mentioned issues.

Other experimental studies aim at assessing visual loss by analyzing the pupillary response to light stimuli^7^. The underlying rationale is that light stimulation within a damage retinal region will elicit reduced pupillary responses because cells involved in this process are malfunctioning or lacking. Most often, the pupil light reflex (PLR) or the post-illumination response (PIPR) characterizing the return to base-line pupil diameter^8^ elicited by brief full-field or focal flashes of light^9^ are used to assess visual field defects^10, 11,12,13^. A caveat is that these tests require dark adaptation for about 10 minutes to evaluate the PLR relative to a well-defined baseline, before numerous PLR separated by about 5 second are collected, as averaging appears necessary to obtain reliable measures^14^. Some studies use different stimuli (e.g. ramping full-field stimuli^15^), but the general outcome is that pupillary activity is perturbed in a number of ophthalmic diseases affecting the retina (hereditary retinitis pigmentosa, diabetic retinopathy…) or the optic nerve (glaucoma, non-glaucomatous optic neuropathies)^16^. Importantly, altered pupillary responses are correlated to structural deficits (RNFL, GCC) measured for instance with Optical Coherence Tomography (OCT)^15,17^. These reflexive physiological objective pupillary responses can be measured non-invasively with little expertise and resources in a short amount of time, dispending individuals to give a subjective, criterion dependent, response.

This approach gained relevance with regards to the discovery of melanopsin containing retinal ganglion cells^18^ and studies of their connectivity and functioning (see ^19,20^ for reviews). Briefly, melanopsin containing retinal ganglion cells project onto the Edinger-Whestphal nucleus through the PON which in turn projects onto the ciliary ganglions that control the iris sphincters^21^. These ganglion cells show slow intrinsic photosensitive activity (ipRGCs) when directly stimulated in the blue range (480 nm), but are also extrinsically activated with faster time constants when stimulated with other wavelengths, through an indirect pathway involving the rods and the cones, as well as bipolar cells^22^. Because these different retinal cells involved in pupil control are selective to different wavelengths, chromatic pupillometry developed in recent years, so as to probe these –intrinsic and extrinsic-circuits independently^23,24,25,26^.

Amongst the pupillary tests used to probe functional deficits, past studies^27^ investigated whether pupil cycle time (PCT) provides a quantitative marker of neuropathies, as in Optic Neuritis for instance. Indeed, PCT is thought to reflect the time constant of the retino-pupillary circuits, which may be lengthened by optic nerve demyelination or by malfunctioning retinal tissue, such as decreased efficacy of photoreceptors transduction, or decreased density of RGCs and ipRGCs. In current practice, periodic cycles of pupillary dilation and constriction are induced by illuminating the pupil margin using a thin beam of a slit-lamp, placed in such a way that pupil size “controls” the amount of light entering the eye: the beam of light first causes a pupil constriction, such that the beam light falls on the iris, outside the pupil, not stimulating the retina; the so induced decreased retinal illumination in turn elicits a pupil dilation, such that the beam light enters the eye again, eliciting a constriction, and so on (Figure 1). With this method, measuring the peak-to-peak time between oscillations revealed that the PCT is significantly longer for patients with Optic Neuritis or Glaucoma ^27,28^.

**Figure 1.**
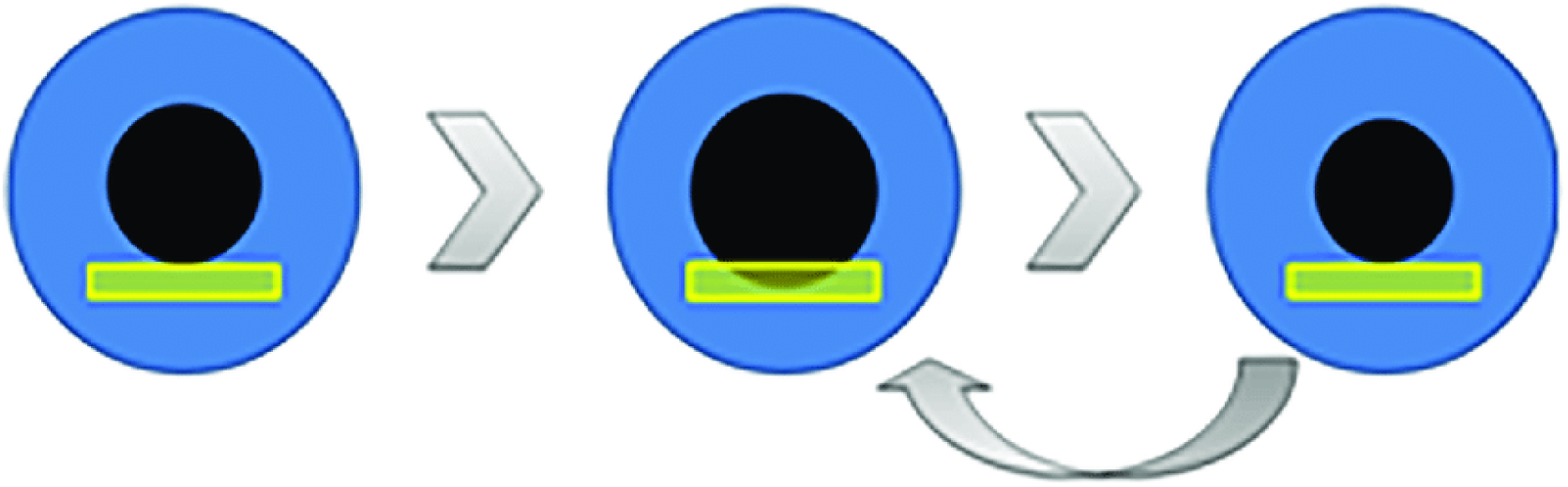
Pupil cycle time elicited by positioning the beam of a slit lamp at the inferior margin of the pupil. With this method, the pupil cycle time is derived from the number of cycles measured using a stop-watch during a fixed period.

**Figure 2:**
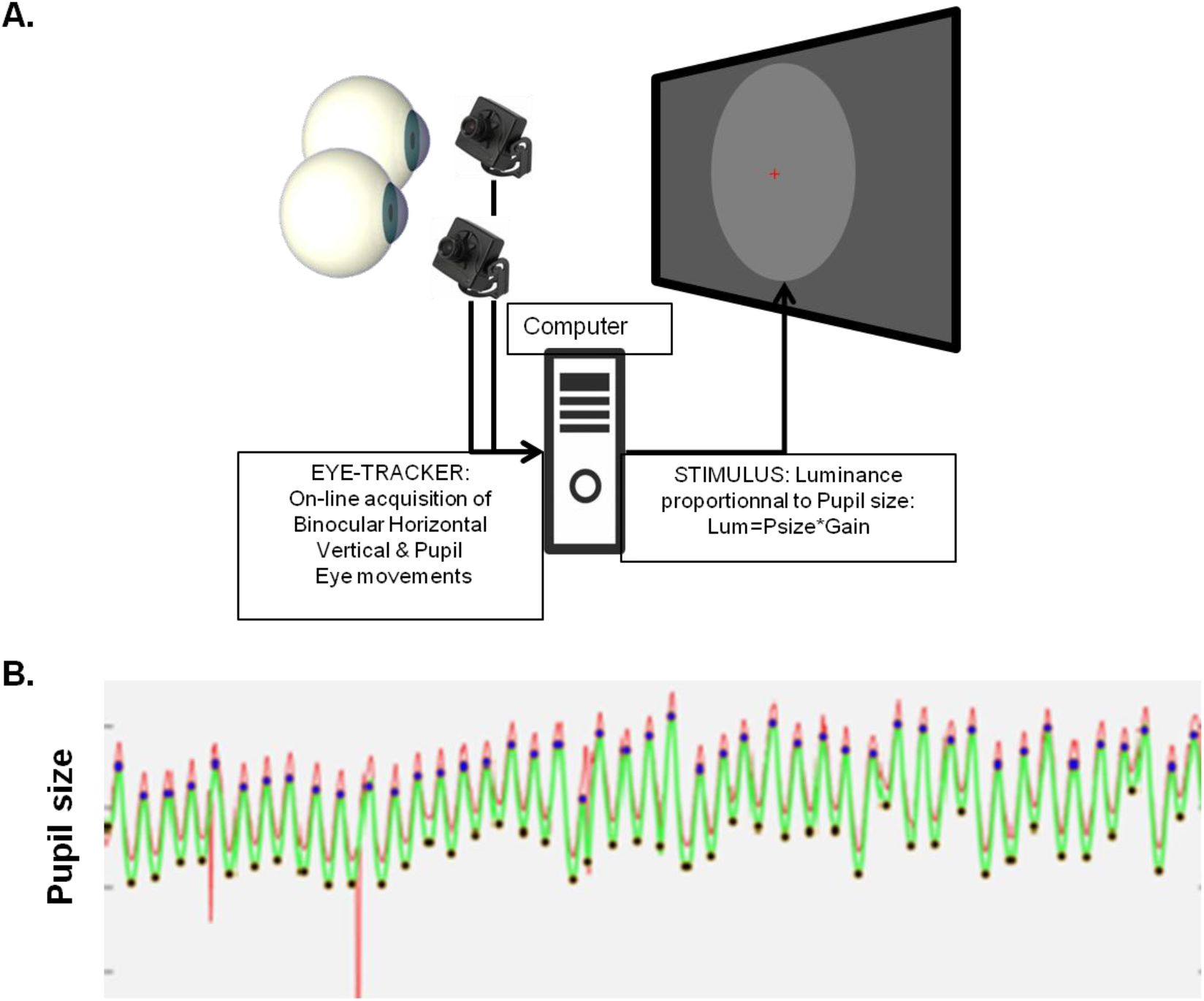
Illustration of the biofeedback settings used to elicit pupillary oscillations. **A**. The diameter of the pupil measured with an eye-tracker is transformed in real-time into a stimulus luminance proportional to pupil size: A large pupil entails a high luminance that elicits a pupil constriction, resulting in decreased stimulus luminance, eliciting in turn a pupil dilation, etc. **B**. Example of 45 seconds of sustained Pupil Oscillation: red trace, raw signal; green trace, corrected signal. Dots show the maxima and minima of PO.

**Figure 3:**
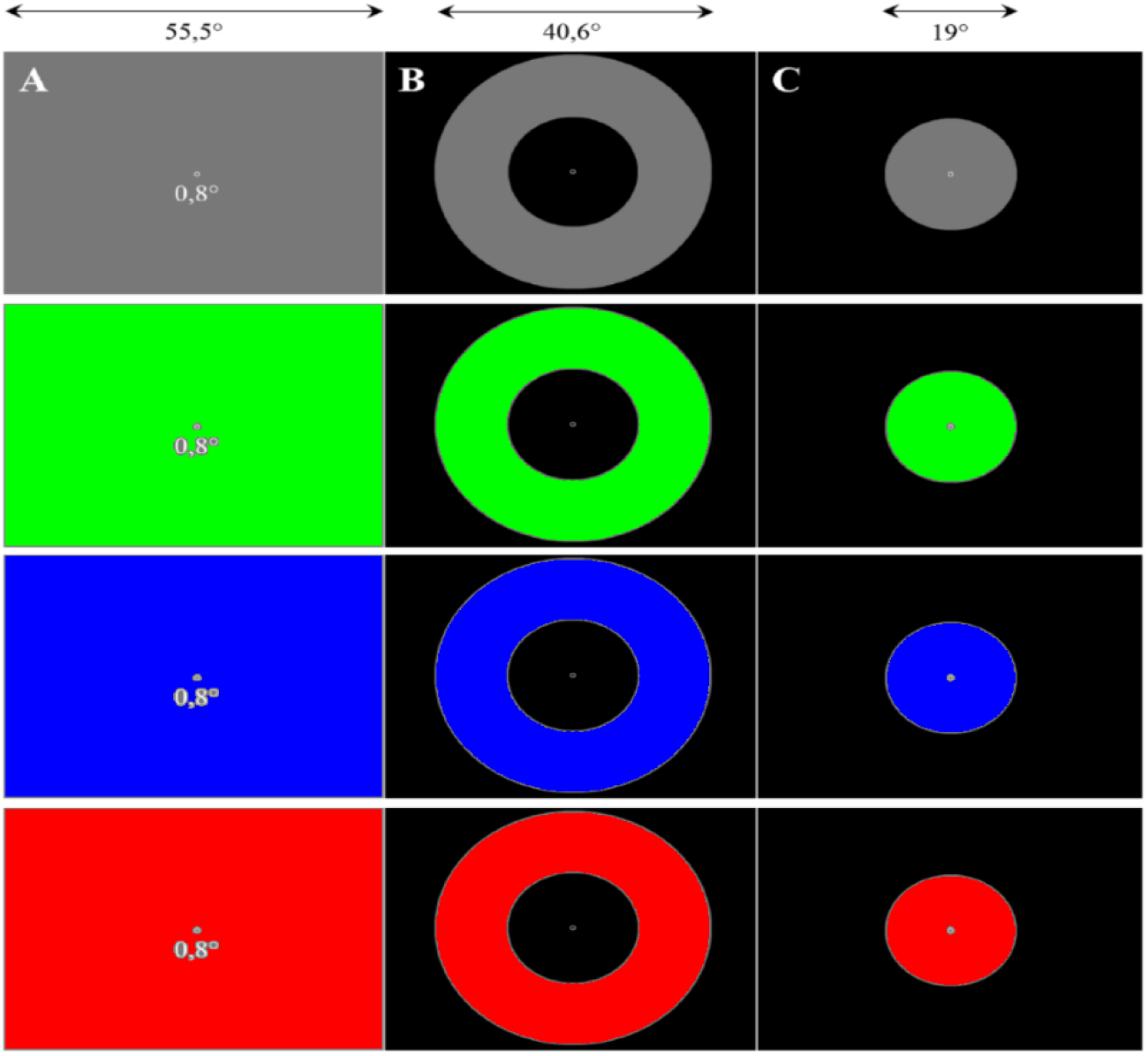
Illustration of the stimuli used in the experiment. Large field, Ring and Disk stimuli with four different colors: grey, green, blue, and red. The large field stimulus was 55.5 × 43°; the outer and inner sizes of the ring stimuli were 40.6 and 19°; the size of the central disk stimuli was 19°.

Despite being short (∼1 minute), the current way of measuring PCT suffers from several limitations:

a. PCT measures require a good practice to master the slit-lamp positioning, and any eye or head movement of the participant may disrupt the pupillary cycling,
b. The slit-lamp stimulus is easily applied onto the iris margin, but not so easily in other locations; hence, it cannot be used to test specific foveal or other retinal regions, or to investigate different stimulus characteristics (for instance stimulus color, size, structure, position or extent). In addition, the size of the driving stimulus –the beam light-on the retina, rather than its luminance, is modulated by pupil size. Therefore, the pupillary cycling itself changes the size of the projection of the beam stimulus that encompasses variable localizations and thus stimulates different retinal regions over time.
c. The outcome measure, the Pupil Cycle time, is measured by counting the number of cycles during a fixed amount of time (using a stop-watch), and by dividing this number by the duration of the test. With this method, only the cycle time (the period of pupillary oscillations) can be measured, without any possibility to quantitatively estimate other characteristics of pupillary oscillations, such as their amplitude, their regularity, the mean pupil size, or the eye-movements and the number of blinks performed during the test.

## Computerized induction of Pupillary Oscillations

Eye-tracking devices coupled to computer based stimulation can overcome these limitations. Lamirel and colleagues (2018) introduced a novel method relying on computerized biofeedback, in which on-line recording of pupil size with an eye-tracker is used to set the luminance of a stimulus displayed on a computer screen in real-time. In this setting, the luminance of a stimulus of arbitrary color, size, and position is set to be proportional to pupil size. Thus, a large pupil entails a high stimulus luminance, which in turn elicits a pupil constriction, resulting in decreased stimulus luminance, and so on. The so-induced pupillary oscillations are recorded, allowing quantitative off-line analyzes to characterize the frequency, the amplitude, the stability of the pupillary oscillations, as well as the eye-movements and number of blinks, providing additional data relevant to characterizing Eye Health, and possibly to distinguishing patients from healthy individuals.

We hypothesized than the PCT characteristics could be altered in both neuropathies and retinopathies, as the retinal circuits involved in pupillary responses are also altered (See e.g. ^17^). We also considered the possibility that in both neuropathies and retinopathies, the integration time of visual stimulation could be lengthened, which could be reflected in the lengthening of the PCT.

In this study, we employed different spatial layouts (Large field stimulation, central disk and peripheral ring) with different colors (Grey, Red, Green, Blue), and applied the method of Lamirel et al. (2018) with patients suffering from Retinitis Pigmentosa (RP), Stargardt disease (SD), Leber Hereditary Optic Neuropathy (LHON), and with healthy individuals (see supplementary Table 1). Retinitis Pigmentosa (RP) is a rare genetic disease characterized by a progressive loss of peripheral vision that can lead to a “tunnel vision” and evolves toward blindness at a late stage; RP is secondary to the death of photoreceptors. Stargardt disease is also a genetic disease, but results in a progressive loss of central vision, due to the accumulation of fatty material on the macula that kills light-sensitive cells. Lastly, LHON, also a rare genetic disease, is an optic neuropathy, affecting central vision, and due to the death of retinal ganglion cells. The choice of these pathologies for this study was dictated by the fact that they affect relatively young individuals, and are thus more rarely accompanied by other comorbidities or by the prescription of drugs that could alter pupil reactivity. So, the choice of these diseases seemed appropriate for this proof of concept study.

**Table 1:** Summary of the ROC analyzes. The left column (Filter) indicates the data set used for the computations. The other columns indicate the pathology, the AUC, the sensitivity, the specificity, the effect size and the number of observations (total and for each pathology). The average, maximum and minimum values are presented in the bottoms rows. Best results are shown in bold character. The table at the bottom presents the results for healthy participants versus data from all patients.

| Filter | PATHO | AUC | Sens | Spec | EffSize | N tot | Npatho | PATHO | AUC | Sens | Spec | EffSize | N tot | Npatho | PATHO | AUC | Sens | Spec | EffSize | N tot | Npatho |
| --- | --- | --- | --- | --- | --- | --- | --- | --- | --- | --- | --- | --- | --- | --- | --- | --- | --- | --- | --- | --- | --- |
| RE+LE All STI All COLORS | RP | 0,85 | 0,74 | 0,83 | 1,09 | 531 | 296 | SD | 0,77 | 0,82 | 0,55 | 0,80 | 503 | 268 | LHON | 0,78 | 0,43 | 0,92 | 0,67 | 360 | 125 |
| RE All STI All COLORS | RP | 0,87 | 0,71 | 0,91 | 1,19 | 269 | 151 | SD | 0,80 | 0,62 | 0,85 | 0,91 | 252 | 134 | LHON | 0,83 | 0,63 | 0,89 | 0,76 | 180 | 62 |
| LE All STI All COLORS | RP | 0,82 | 0,83 | 0,68 | 0,98 | 262 | 145 | SD | 0,74 | 0,87 | 0,51 | 0,68 | 251 | 134 | LHON | 0,75 | 0,40 | 0,90 | 0,57 | 180 | 63 |
| RE+LE All STI Col Red | RP | 0,85 | 0,76 | 0,89 | 0,65 | 100 | 46 | SD | 0,81 | 0,65 | 0,93 | 0,92 | 108 | 54 | LHON | 0,82 | 0,48 | 0,98 | 0,62 | 81 | 27 |
| RE+LE All STI Col Green | RP | 0,89 | 0,83 | 0,77 | 1,34 | 186 | 113 | SD | 0,82 | 0,79 | 0,73 | 0,80 | 167 | 94 | LHON | 0,85 | 0,53 | 0,97 | 0,57 | 113 | 40 |
| RE+LE All STI Col Blue | RP | 0,86 | 0,86 | 0,78 | 1,01 | 111 | 57 | SD | 0,81 | 0,82 | 0,69 | 0,82 | 114 | 60 | LHON | 0,83 | 0,68 | 0,87 | 0,95 | 82 | 28 |
| RE+LE All STI Col Grey | RP | 0,86 | 0,86 | 0,72 | 1,24 | 134 | 80 | SD | 0,75 | 0,92 | 0,44 | 0,77 | 114 | 60 | LHON | 0,84 | 0,70 | 0,85 | 0,65 | 84 | 30 |
| RE+LE Center All Colors | RP | 0,76 | 0,72 | 0,71 | 0,66 | 159 | 86 | SD | 0,75 | 0,74 | 0,67 | 0,63 | 155 | 82 | LHON | 0,76 | 0,51 | 0,84 | 0,38 | 112 | 39 |
| RE+LE Ring All Colors | RP | 0,91 | 0,87 | 0,81 | 1,33 | 161 | 89 | SD | 0,80 | 0,84 | 0,61 | 0,88 | 154 | 82 | LHON | 0,87 | 0,68 | 0,89 | 0,93 | 110 | 38 |
| RE+LE Full All Colors | RP | 0,88 | 0,80 | 0,83 | 1,27 | 211 | 121 | SD | 0,81 | 0,77 | 0,68 | 0,89 | 194 | 104 | LHON | 0,81 | 0,48 | 0,92 | 0,71 | 138 | 48 |
| RE+LE Center Col: Red | RP | 0,81 | 0,87 | 0,72 | 0,07 | 33 | 15 | SD | 0,85 | 0,94 | 0,67 | 0,43 | 36 | 18 | LHON | 0,75 | 0,67 | 0,94 | 0,02 | 27 | 9 |
| RE+LE Center Col: Green | RP | 0,85 | 0,68 | 0,95 | 0,78 | 47 | 28 | SD | 0,90 | 1,00 | 0,68 | 0,63 | 43 | 24 | LHON | 1,00 | 1,00 | 1,00 | 0,30 | 29 | 10 |
| RE+LE Center Col: Blue | RP | 0,92 | 0,88 | 0,89 | 0,69 | 35 | 17 | SD | 0,80 | 0,85 | 0,78 | 0,83 | 38 | 20 | LHON | 0,79 | 0,70 | 0,83 | 0,56 | 28 | 10 |
| RE+LE Center Col: Grey | RP | 0,79 | 0,73 | 0,83 | 0,88 | 44 | 26 | SD | 0,76 | 0,65 | 0,78 | 0,53 | 38 | 20 | LHON | 0,94 | 0,90 | 0,89 | 0,43 | 28 | 10 |
| RE+LE Ring Col: Red | RP | 1,00 | 1,00 | 1,00 | 1,07 | 33 | 15 | SD | 0,95 | 0,89 | 1,00 | 0,90 | 36 | 18 | LHON | 1,00 | 1,00 | 1,00 | 0,91 | 27 | 9 |
| RE+LE Ring Col: Green | RP | 0,94 | 0,86 | 0,89 | 1,56 | 47 | 29 | SD | 0,85 | 0,96 | 0,67 | 0,95 | 42 | 24 | LHON | 1,00 | 1,00 | 1,00 | 0,69 | 28 | 10 |
| RE+LE Ring Col: Blue | RP | 0,96 | 0,95 | 0,83 | 1,02 | 37 | 19 | SD | 0,83 | 0,65 | 0,94 | 0,64 | 38 | 20 | LHON | 0,90 | 0,56 | 1,00 | 0,88 | 27 | 9 |
| RE+LE Ring Col: Grey | RP | 0,90 | 0,88 | 0,78 | 1,39 | 44 | 26 | SD | 0,89 | 0,90 | 0,78 | 0,90 | 38 | 20 | LHON | 1,00 | 1,00 | 1,00 | 0,96 | 28 | 10 |
| RE+LE Full Col: Red | RP | 0,95 | 0,81 | 0,94 | 0,71 | 34 | 16 | SD | 0,89 | 0,89 | 0,72 | 1,30 | 36 | 18 | LHON | 1,00 | 1,00 | 1,00 | 0,75 | 27 | 9 |
| RE+LE Full Col: Green | RP | 0,91 | 0,91 | 0,75 | 1,49 | 92 | 56 | SD | 0,84 | 0,76 | 0,78 | 0,81 | 82 | 46 | LHON | 0,87 | 0,85 | 0,78 | 0,62 | 56 | 20 |
| RE+LE Full Col: Blue | RP | 0,91 | 1,00 | 0,67 | 1,18 | 39 | 21 | SD | 0,96 | 0,95 | 0,89 | 0,91 | 38 | 20 | LHON | 0,90 | 0,67 | 1,00 | 1,18 | 27 | 9 |
| RE+LE Full Col: Grey | RP | 0,99 | 1,00 | 0,89 | 1,25 | 46 | 28 | SD | 0,73 | 0,65 | 0,78 | 0,74 | 38 | 20 | LHON | 0,96 | 0,90 | 0,94 | 0,38 | 28 | 10 |
| Average | RP | 0,89 | 0,84 | 0,82 | 1,04 |  |  | SD | 0,82 | 0,81 | 0,73 | 0,80 |  |  | LHON | 0,88 | 0,72 | 0,93 | 0,66 |  |  |
| Maximum |  | 1,00 | 1,00 | 1,00 | 1,56 |  |  |  | 0,96 | 1,00 | 1,00 | 1,30 |  |  |  | 1,00 | 1,00 | 1,00 | 1,18 |  |  |
| Minimum |  | 0,76 | 0,68 | 0,67 | 0,07 |  |  |  | 0,73 | 0,62 | 0,44 | 0,43 |  |  |  | 0,75 | 0,40 | 0,78 | 0,02 |  |  |

| HP versus all Patients |  |  |  |  |  |  |
| --- | --- | --- | --- | --- | --- | --- |
|  | AUC | Sens | Spec | EffSize | N tot | Npatho |
| RE+LE All STI All COLORS | 0,78 | 0,97 | 0,28 | 0,95 | 924 | 689 |
| RE All STI All COLORS | 0,81 | 0,95 | 0,35 | 1,06 | 465 | 347 |
| LE All STI All COLORS | 0,75 | 0,97 | 0,28 | 0,83 | 459 | 342 |
| RE+LE Ring Col: Red | 0,92 | 0,88 | 0,83 | 1,07 | 60 | 42 |
| RE+LE Ring Col: Green | 0,86 | 1,00 | 0,39 | 1,30 | 81 | 63 |

We here tested whether PCT induced by stimuli with different spatial configurations and chromaticity could probe the integrity of the retinal circuits in the above mentioned pathologies. The goal of this study was to evaluate the capability of our computerized biofeedback setting to reliably induce pupillary oscillations, and to provide a novel data set for the pathologies studied herein. We hypothesized that PCT characteristics could be altered, in accordance with the known damaged regions and defective cells proper to each of these pathologies.

## Material and Method

### Participants

Fourteen patients with Retinitis Pigmentosa (RP: mean age: 41, sd: 11; 6 women), 14 patients with Stargardt disease (SD: mean age: 38, sd: 9; 5 women), 9 patients with Leber hereditary optic neuropathy (LHON: mean age: 33, sd: 7; 4 women) and 14 healthy participants (HP: mean age: 37, sd: 10; 6 women) were included in the study. All participants were aged between 20 and 58 years. The inclusion criteria for the healthy volunteers were a binocular corrected visual acuity greater or equal to 8/10 (≤ 0.1 logMAR) and a normal visual field (Supplementary table 1). Patients and healthy participants were tested without corrections (lens or glasses) during the main experiment.

### Protocol

All participants passed a full ophthalmic assessment, including visual acuity (ETDRS right and left eye, and binocular), color vision test (saturated and desaturated 15 Hue tests: D-15d), visual field test (Humphrey Field Analyzers, HFA, model Octopus 900, using isopter V4 to III), contrast sensitivity (Pelli-Robson, left, right eye and binocular), fundus examination as well as a macular (macular volume and thickness) and optic nerve (peripapillary retinal nerve fiber layer thickness) optical coherence tomography (OCT, Spectralis Heidelberg, Engineering, Heidelberg, Germany). Whenever possible, the ophthalmic assessment was performed on the same day as the pupillary tests.

### Apparatus

The stimuli were back-projected at 60 Hz using a Titan 1080p video projector (Digital Projection, Ldt) on a translucent screen (55.5 × 41.63 degrees of visual angle). Participants were conformably installed at 128 cm from the screen, while resting their head on a chin-rest. Pupil modulations and eye-movements of both eyes were recorded with an eye-tracker (EyeLink II, 500 frames/s., SR Research, Ldt), and down-sampled to 60 frames/s. To avoid the burden of wearing a helmet for a long time, the 2 cameras of the eye-tracker were deported and fixed on the chin-rest. The cameras were adjusted for each subject, so as to obtain a good image of both pupils. Each eye could be stimulated independently, using a large mobile black cardboard that masked the display screen to one eye. Dedicated custom software (Jeda) was used for stimulus generation and eye-movement recordings.

### Stimuli

Three different stimuli –a central disk, a peripheral ring and a large field covering the entire screen were used. The central disk stimulus covered 19° of visual angle. The inner and outer borders of the ring stimulus were 19° and 40° respectively. These stimuli could have different “colors” (Red, Green, Blue and Grey), corresponding to modulations of the different RGB guns of the display. The grey stimulus was obtained by modulating all RGB guns at once. The stimulus luminance was updated on every frame (at 60 Hz), depending on pupil size, at a rate much faster than pupillary responses. Conversion of pupil size into stimulus luminance was achieved by using 2 parameters: a gain, G (multiplicative factor), and an offset, O. The equation used for this conversion was of the form:

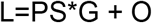

with L corresponding to the stimulus intensity (value between 0 and 255 applied to the RGB display guns), and PS corresponding to the size of the pupil delivered on-line by an eye-tracker. When necessary, G and O were adapted to participants with whom the default parameters did not reliably induce pupillary oscillations. In some cases, induction of oscillations failed, and the corresponding conditions were skipped, resulting in imbalanced number of trials for each condition and patients’ groups.

The amplitude of luminance modulations was proportional to pupil size, and could in principle range from 1 cd/m^2^ to 181.10 cd/m^2^ (min and max of display luminance). In practice, the maximum and minimum luminances depended on the subjects, but were much less that the display capabilities. Each of the 12 PCT tests lasted 45 seconds.

### Procedure

The session started with positioning the participants and adjusting the cameras of the eye-tracker. After 10 minutes of adaptation to the low ambient light of the testing room, during which a questionnaire was given to the participants, a 5 point eye calibration procedure was given before series of short (<1 minutes) tests, including PLR measures, RAPD assessment, as well as other tests, including the large-field and quadrant Multiple Pupillary Frequency Tagging tests (mPFT, see Ajasse et al, 2022). Overall, a session lasted about 2 hours. PCT inductions were part of these tests, for which participants were simply asked to maintain fixation at the center of the screen, marked by a small white circular fixation ring, with no other concurrent task. A brief rest, adapted to each participant, separated the different runs, and was used when necessary to change the stimulated eye (Right or Left; note that eye-movements and pupillary responses of both eyes were always recorded during a run). Pupil oscillations were recorded for all 12 stimuli, in monocular conditions (Right and Left eyes). This design results in 24 conditions per session per participant, with overall 1224 expected recordings (2 eyes x 3 spatial configurations x 4 colors x 51 participants). However, some participants did not perform in all conditions, due to fatigue or to technical issues, as for instance inappropriate parameters used to convert pupil size into stimulus luminance. In addition, some participants made many blinks or eye-movements that perturbed pupillary recordings. As a consequence, 1023 trials were recorded. 44 trials could not be analyzed because of a missing or very noisy pupillary signal, generally related to a very large number of blinks, such that 979 trials were included in the analysis (465 and 514 for the right and left eye, respectively).

### Data analyzes

Analyzes were performed with Matlab R2018b (The MathWorks, Natick, MA, USA). The raw pupil data were down-sampled at 60 Hz and corrected for blinks and artifacts. Blinks and spurious data were detected on the pupillary data using both a velocity and an acceleration threshold, chosen once for the whole data set. Lacking or spurious data were replaced by a smoothed linear interpolation, using a pre- and post-blink offset of 4 samples (66.6 msec). As the initial and final pupillary responses were often noisy, the signal was trimmed by 120 samples (3 sec.), so that 39 seconds of pupillary activity were used for the analyses. The corrected pupillary signals were then z-scored, and a Fast Fourier transform (FFT) was computed on the so-corrected pupillary signals, together with a time frequency analysis. From these analyzes, we derived several variables, including the Oscillation Frequency (OF, the inverse of the Pupil Cycle Time) and the Power at the dominant frequency (OP). We also computed the mean FFT power in the range 0.5-2Hz. Using the z-scored pupillary recordings, we further computed the averaged peak-to-trough amplitude of oscillations (OA), the standard deviation of their distribution (OAv), the averaged peak-to-peak durations (OD, corresponding to the period of the PO), the standard deviation of their distribution (ODv), the slope of the overall pupillary response computed over 39 seconds (the “pupillary escape” observed during a trial), the mean vertical and horizontal eye positions, as well as the number of corrected data (corresponding to blinks and spurious data). We compared these variables across and within the different groups of participants, to identify which variables were most relevant biomarkers of the diseases at stake. To that aim, we first computed analyses of variance using the Matlab *anovan* function with each of the variables described above.

We then used some, or all, of these variables to compute the area under the receiving operating characteristic curves (AUC of ROC), so as to evaluate the extent to which these variables allow discriminating patients from healthy participants. The area under the curve (AUC) and the corresponding sensitivity, Ss, and specificity, Sp, were computed using the *fitglm* and *perfcurve* Matlab functions: first, a logistic regression model was computed with the Matlab *fitglm* function using the variables listed above. Second, the probability distribution returned by the *fitglm* function as scores was used to compute the ROC curves with the Matlab function *perfcurve*. ROC curves were computed separately for each eye, each stimulus configuration and each color, or for pooled colors or pooled stimuli or both, to evaluate which variables allowed the best discrimination of the different patient groups (SD, RP and LHON) from healthy participants (HP).

## Results

Figure 4 shows the different steps of single trial analysis (Additional examples are available in Supplementary figure 1). Eye-movements and pupil activity where first checked visually (Figure 4a), before analyzing the eye positions and pupil oscillations over time (Figure 4b). Pupil data were then corrected for blinks and artifacts, and spurious data were replaced by a smoothed linear interpolation (Figure 4c). We then computed a linear fit on the corrected pupillary response to determine the overall slope of PO, before a Fourier transform was applied to the corrected signal to extract the dominant frequency in the range 0.5-2 Hz (Figure 4d). Relevant variables (see methods) were finally computed for later classification tests.

**Figure 4:**
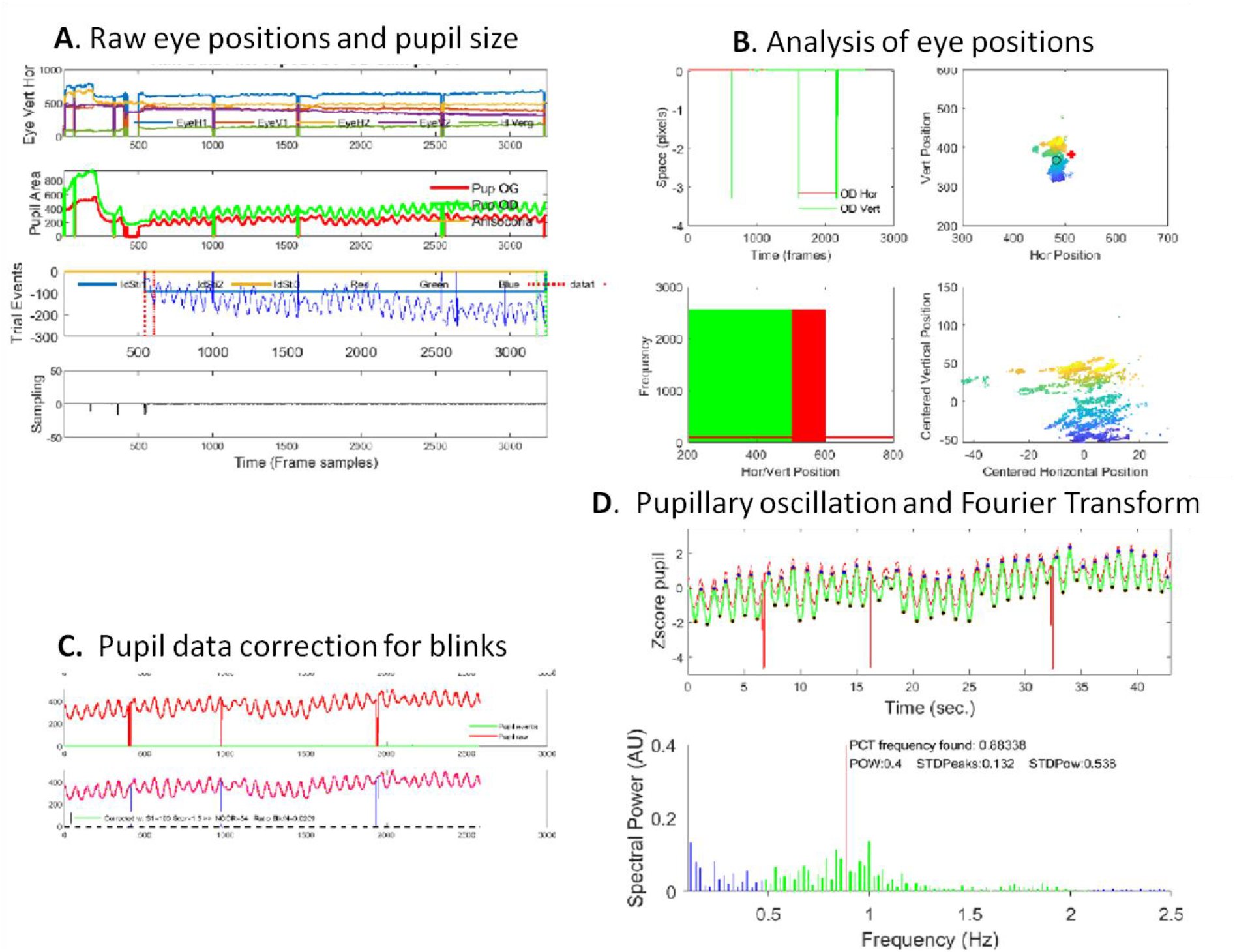
Example of the pipe line analysis applied to each trial. **A**: Visual check of raw eye-positions and pupil size. **B**: Analysis of vertical and horizontal eye-movements during a trial. **C**: Correction of blinks and spurious pupil data, replaced by a smooth linear interpolation. **D**: Analyses performed on the corrected pupil data. Top panel: Pupil oscillations over time (raw pupil size: red line; green: corrected pupil size); blue disks show the peaks of pupillary oscillations, from which the amplitude and period of pupillary oscillations were computed, together with their variability. Bottom: Fourier transform applied on the corrected pupil data. The maximum power (OP) in the range 0.5-2 Hz was taken as the pupillary oscillation frequency (OF).

In several recordings, the eye-positions were missing or were offset relative to the fixation point, because performing a good calibration was difficult with some patients, and because eye drift during or across runs could occur. However, accurate eye-calibration was not required in this study, as pupil size measurements do not rely or depend upon calibration accuracy.

Group analyzes were then conducted to compare the effects of the variables derived from each trial, for the 12 conditions tested for each eye (4 “colors” x 3 spatial configurations). These variables include the oscillation frequency (OF) and the oscillation power (OP) derived from the FFT. We also analyzed the oscillation amplitude (OA) and its variability (OAv), the oscillation period (OD) and its variability (ODv).

The distributions of these measures are shown in Figure 5, 6 and 7. Each figure shows the results for the one spatial configuration (Large field, Peripheral Ring and Center disk). Within each figure, the different panels show the values of one variable for the 4 different colors and the 4 patient groups.

**Figure 5:**
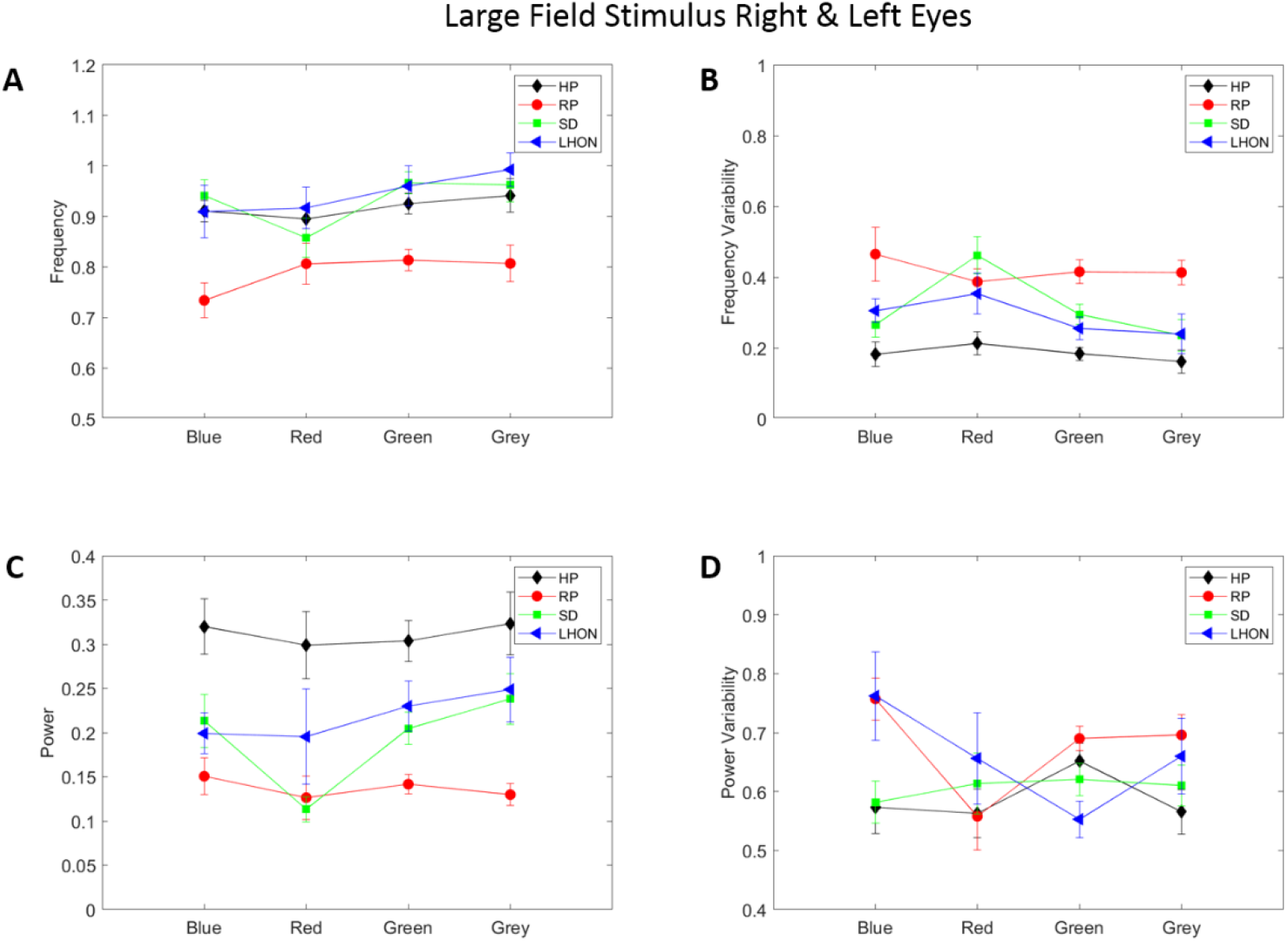
Characteristic measures of POs for the <u>Large Field stimulus</u>, for the pooled Right and Left eye data. **A**. Distribution of Pupillary Oscillation Frequency as a function of the stimulus Color, for the 4 groups: HP: black symbols, Healthy Participants; RP: red symbols, Retinitis Pigmentosa; SD : green symbols, Stargardt disease; LHON : blue symbols, Leber Hereditary Optic Neuropathy. **B**. Oscillation frequency variability. Symbols as in A. **C** Pupillary Oscillation Power, Symbols as in A. **D** Variability of Pupillary Oscillation amplitude. Symbols as in A. Error bars represent one standard error.

**Figure 6:**
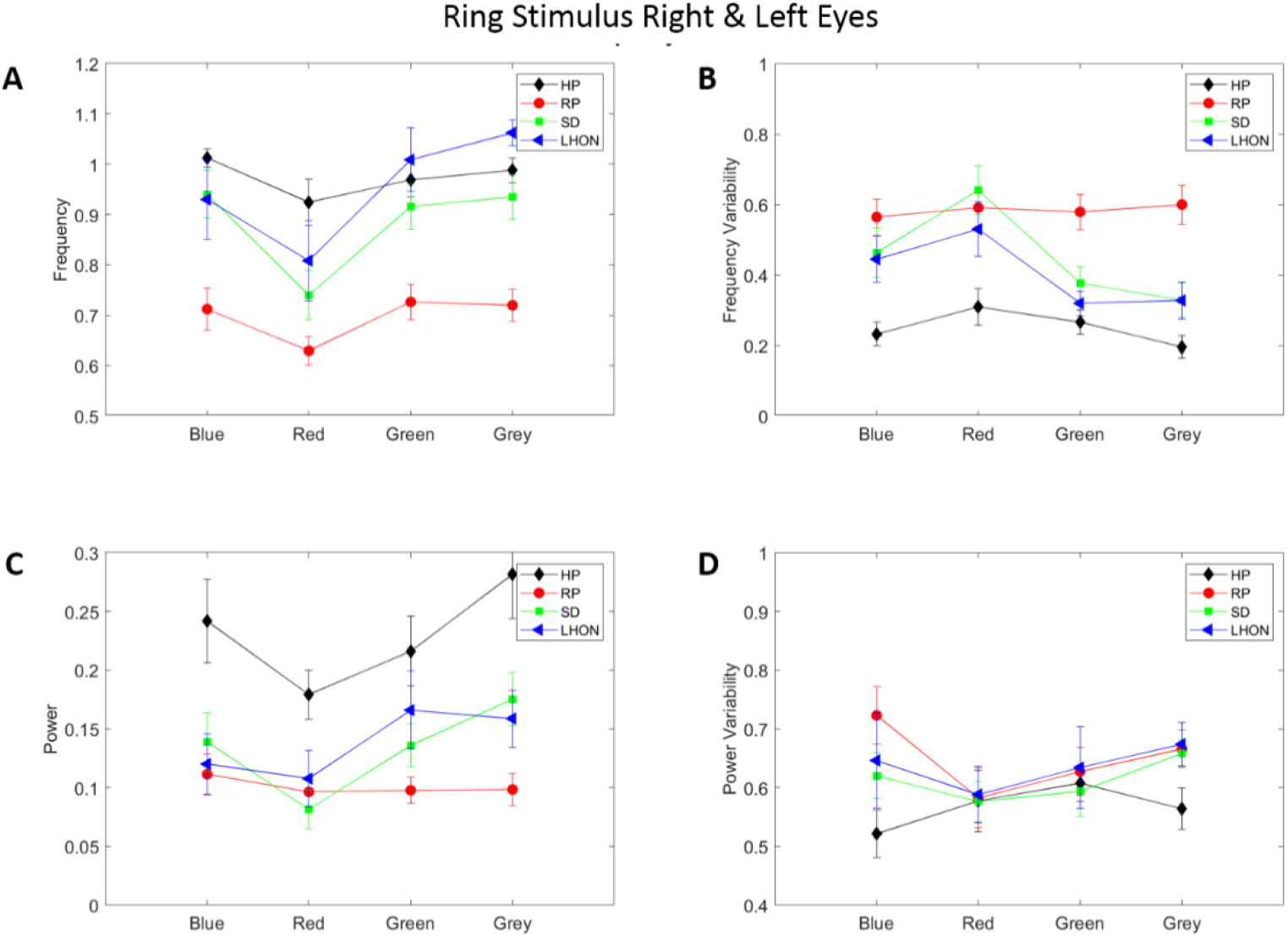
Characteristic measures of PO for the <u>Ring stimulus</u>, for the Right and Left eyes. **A, B, C, D** same as in Figure 5.

**Figure 7:**
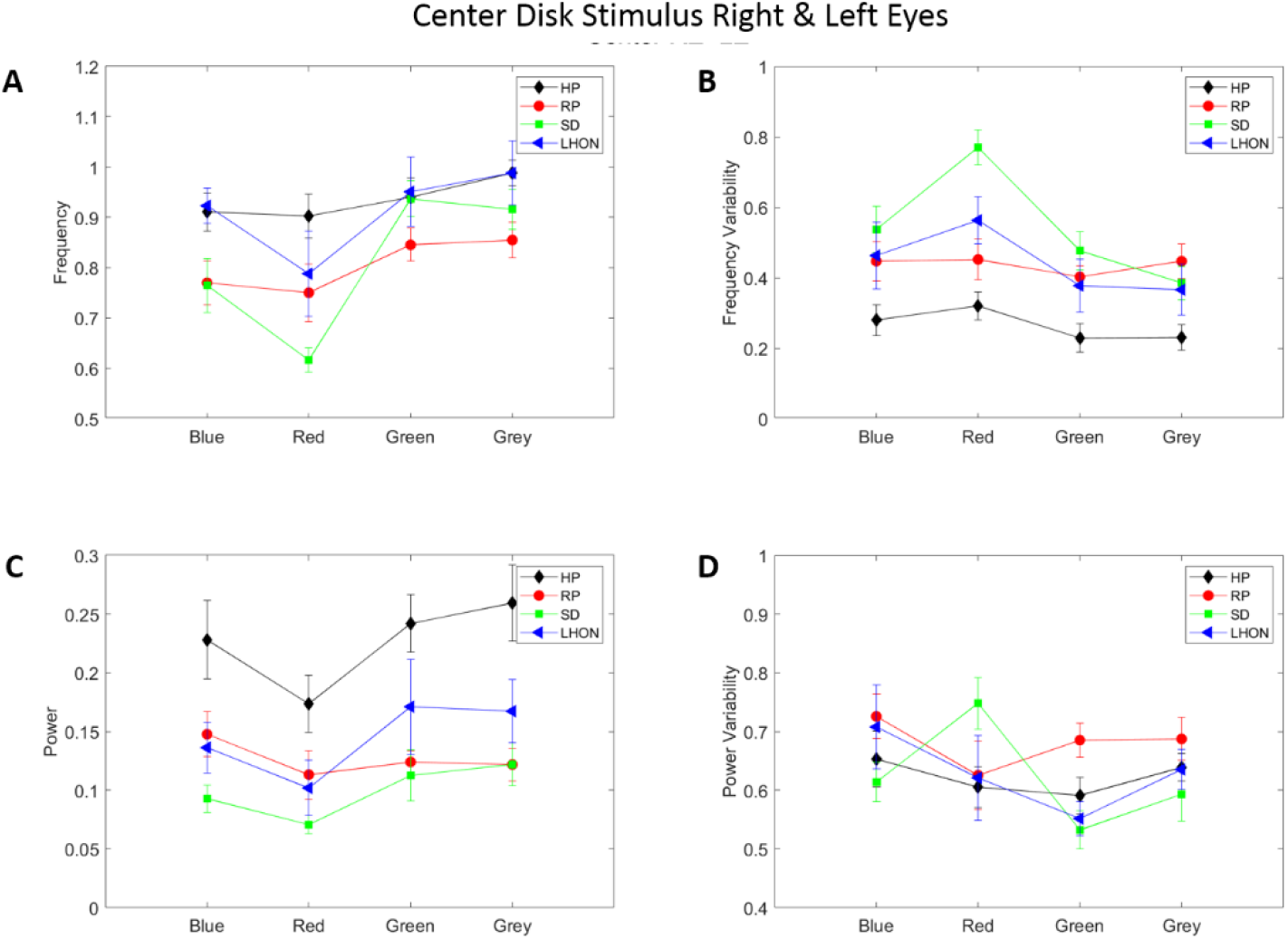
Characteristic measures of PO for the <u>Center disk stimulus</u>, for the Right and Left eyes. **A, B, C, D** same as in Figure 5.

As it can be seen in Figures 5, 6 and 7 the variables derived from the pupillary recordings depend on the spatial layout (Large field, Ring or Center disk), the stimulus color (Blue, green, red or grey) and differ for the different groups. An analysis of variance computed on the whole data set indicates that the Frequency, the Frequency Variability, and the Oscillation Power differed significantly as a function of the spatial configuration and of the color for the different groups (all p<0.0001, see Supplementary table 2). Despite other variables and interactions between variables sometimes showing significant effects, we do not present all results in details herein (all statistical results are available in Supplementary Table 2). We here focus on the Oscillation Frequency, the Frequency Variability and the Oscillation Power. Notably, we found that the oscillation power is larger and the frequency variability is smaller for the Healthy participants as compared to the patient groups in all conditions. Note that these variables cannot be computed with the usual slit-lamp setting (see introduction), and are therefore never considered for the functional assessment of visual defects.

### Effects of Spatial Configuration

Overall, the Large field stimuli induce oscillations of greater power than the other spatial configurations. This is expected as these large stimuli recruit a larger region of the visual field than the other spatial layouts. This large field stimuli do not, however, entail important changes in the Oscillation frequency, but do reduce the Frequency variability, as compared to the Ring and Disk stimuli (compare for instance the Frequency variability of the healthy participants for the 3 spatial configurations).

As expected, the effects of the spatial layout are mostly pronounced for the patient groups, who present defects in disease dependent regions of the visual field. As a matter of fact, these defective regions decrease their potential contribution to pupillary activity, which can partly account for the differences between healthy participants and patients, as fewer cells are contributing to the pupillary activity. We now detail some of the results with regards to the stimuli characteristics and the diseases’ specificities.

For the RP group, the PCT variables differ most from healthy participants with the Large field and Ring stimulus, but less so with the Central disk stimulus, in accordance with the peripheral damage proper to this disease. However, inducing Pupillary Oscillation appears overall difficult with RP patients, even with a central stimulation. Conversely, SD and LHON patients show larger differences with healthy participants with the Ring and Central disk stimuli. These differences do not concern the Oscillation Frequency, which remains comparable for the 3 spatial layouts in HP, SD and LHON (with the exception of red stimuli, see below), but rather the Oscillation Power which is lower for SD and LHON patients as compared to healthy participants, and the Frequency Variability that progressively increases with the Ring and with the Center disk (compare the Frequency Variability of SD and LHON patients in figures 5, 6 and 7).

### Effects of Stimulus Color

Although the effects of color on PCT variables are overall modest, Red stimuli elicit oscillations of lower frequency and lower amplitude than the other colors in most participants. This is presumably related to the lower intrinsic luminance of these red stimuli as compared to the green and grey stimuli, as well as to the lower efficacy of red stimuli in inducing pupillary oscillations, as compared to blue stimuli that, despite their low luminance, directly target ipRGCs that efficiently drive the pupil (see introduction). Interestingly, the results for the red stimuli are worse in SD patients as compared to the other groups, especially for the central disk stimulus. This finding is in line with the known alteration of color vision due to the damages to retinal cones in this disease^30^. Note that green stimuli do however reliably entrain pupillary oscillations. The difference between red and green stimuli in SD patients is reminiscent of an imbalance between L-cone and M-cone reported elsewhere^31^, and add evidence that L-cones may be specifically at stake in this disease.

A similar trend is observed for LHON patients with the central disk stimulus, who however do not present a similar pattern for the Large field and Ring stimuli, as the imbalance between PO variables with red and green stimuli is less than that found in SD patients, and more similar to that observed in healthy participants. Finally, we observe that PCT variables in RP patients are not very different for red and green stimuli, and are similar to those of healthy participants, despite showing overall alterations for all the PCT variables considered herein.

### Identifying patients using PCT variables

To determine the extent to which the variables derived from the pupillary oscillations allow classifying patients from healthy subjects, we conducted several ROC analyzes. To that aim, we used the variables that best characterize the PO (see above), but also considered the multidimensional aspect of a participant behavior during a trial, including blinks (approximated by the ratio of the number of corrected data to the number of recorded data) and other pupillary characteristics (e.g. the slope fitted to pupillary signal, reminiscent of the “pupillary escape” described in the literature for long lasting stimulations).

We first conducted these ROC analyzes using all available data, pooling data from the two eyes and the different spatial layouts and colors, and then selected specific data sets, differentiating the two eyes or the different experimental conditions. The summary of these analyzes are shown in Table 1.

Overall, ROC analyzes reflect the results of figures 5, 6 and 7. The variables characterizing POs allow classifying patients form healthy participant with good to excellent performance, depending on the data sets used to compute AUC, sensitivity and specificity.

The effect size (Hedge’s score) for each comparison is large on average (mean effect size: RP=1.04, SD=0.8, LHON=0.66), although these values depend on the data sets.

We also computed AUC of ROC contrasting the data of healthy participants to all the data from the patients (Table 1, bottom). These analyzes show the AUCs are moderate, but that sensitivity is high, while specificity is low, as expected when pooling data from different diseases.

From this synthetic table, it appears that the stimulus configurations yielding the best classification results correspond to a red or green Ring stimulus.

## Discussion

We tested a biofeedback setting able to induce pupillary oscillations with different stimulus configurations in healthy participants and patients with retinopathy or neuropathy. We found that this method was very efficient in eliciting pupillary oscillations, from which we could extract relevant information that allowed classifying patients from healthy participants with good accuracy. In particular, we found that Red and Green Ring stimuli provide the best configuration to sort patients from healthy subjects.

The frequency of PO elicited with our method is comparable to that obtained with the usual way of measuring PCT. Although PCT has mostly been used in patients with optic nerve damage (e.g. Optic neuritis) and considered as reflecting the slowing down of the conduction of retinal responses through the –demyelinated-optic nerve, the present data suggest PCT measures can also be affected in diseases not primarily concerned with optic nerve malfunction. Our findings suggest that damage to the retinal tissue, whether it is photoreceptors as in RP and SD, or ganglion cells as in LHON, possibly including ipRGCs, affect pupillary oscillations, presumably by altering the inner retinal circuits in charge of driving the pupil.

The short duration of the test and the information it brings about the functioning of the retino-pupillary circuits suggest it could be part of the functional assessment needed to make a diagnosis or to follow-up the evolution of a disease. Note that the test duration could be shortened further, as pupillary oscillations are stable over time, as least in healthy participants. Analyzing POs in real time would permit detecting when the characteristics of pupillary oscillations reach those of a healthy standard, which would permit to adjust the duration of the test depending on the on-line measure of relevant PO parameters.

Finally, note that in an another study on healthy participants, we observed that the characteristics of Pupillary Oscillations do not appear to depend upon the duration of dark adaptation before the test, suggesting that PCT measures could be performed without this long and tedious dark adaptation phase that limits its usability in routine clinical assessment of Eye Heath. Further note that in the present study, PCT measures were part of a long lasting session of about 2 hours, however, PCT measures for the 24 tests performed at varying times were comparable in healthy participants, suggesting that PCT characteristics can be evaluated without requiring constraining conditions.

### Limitations of the study

This study presents several limitations. First, the small number of participants, and the fact that the diseases considered in this study are rare, and mainly affect young adults, prevents firm conclusions about the interest of PCT measures for the functional assessment of vision in other pathologies. Second, in the perspective of using POs to assess functional defects, the number of conditions tested herein is much too large and take much too long, and should thus be limited to the most relevant conditions (i.e. Red and/or Green Ring), before it can be routinely used as an additional tool for assessing visual functions. Another issue is the lack of a precise control of the “colors” used here, as no attempt to use specific wavelengths so as to target specific cells (ipRGCs, rods and cones) was made. Although this can appear problematic if the objective is to uncover the precise organization of the damaged circuits in different pathologies, this presents the advantage of not requiring sophisticated, well controlled, –and possibly expensive-devices. Finally, one can question the choice of the spatial configurations used herein, especially the size of the disk and ring stimuli, that could be adapted to better stimulate specific retinal regions.

This proof of concept should be enlarged to more common diseases to ensure it provides relevant and reliable biomarkers of other pathologies. In this view, preliminary results with glaucoma patients, to be presented elsewhere, indicates that PCT characteristics are also altered in this disease, in line with other studies^29^.

## Conclusions

Using the biofeedback settings introduced by Lamirel et al. (2018) with patients suffering from optic neuritis proved feasible with other pathologies. This method brings a number of benefits when compared to the usual way of measuring the PCT with a slit-lamp. In particular, recording pupillary activity with an eye-tracker allows computing several variables that would not be accessible otherwise, such as the power or the variability of pupil oscillations over time. In the present study, these variables were found to significantly differ depending on the disease at stake, while the frequency of pupillary oscillations (e.g. the inverse of the period of pupillary oscillations usually estimated in current practice) is sometimes similar and does not always allow distinguishing patients from healthy subjects. In addition, the finding that the spatial stimulus configuration and color elicit pupillary oscillations with disease dependent characteristics can provide insights into the physiopathology of these diseases.

Finally, the induction of pupillary oscillations with this method is simple and fast (45 seconds) and could thus be used as an additional functional test to characterize, and possibly diagnose and follow-up, different pathologies.

## Data Availability

All data produced in the present study are available upon reasonable request to the authors

## Acknowledgments

The authors are very grateful to all the participants who accepted to contribute to this study. The study was funded by the “Fondation Voir & Entendre”, and took place in the “Street Lab” facilities at the “Institut de la Vision”.

## Author contributions

JL and CV wrote the manuscript; JL wrote software for visual stimulation and data acquisition; SA, CC and JL defined the protocol; SA and JL analyzed the data; CV and CC conducted the patients’ examination; MS and CR managed the recruitment of the patients.

## Ethics declarations

All methods were carried out in accordance with relevant guidelines and regulations. All experimental protocols were approved by an institutional committee. The protocol conformed to the Declaration of Helsinki and was approved by the French local ethic committee (“Comité de Protection des Personnes Ile de France XI”, Ref CPP: 18002, Code: P1705, IDRCB: 2017-A03236-47). All participants gave their written informed consent to participate in the study, and were free to leave the study whenever they decided to do so.

