## Supplementary Figures and Tables for "Computerized biofeedback to characterize Pupil Cycle Time (PCT) in neuropathies and retinopathies"

### Supplementary material

Supplementary Figure 1: Examples of pupillary oscillations for different participants

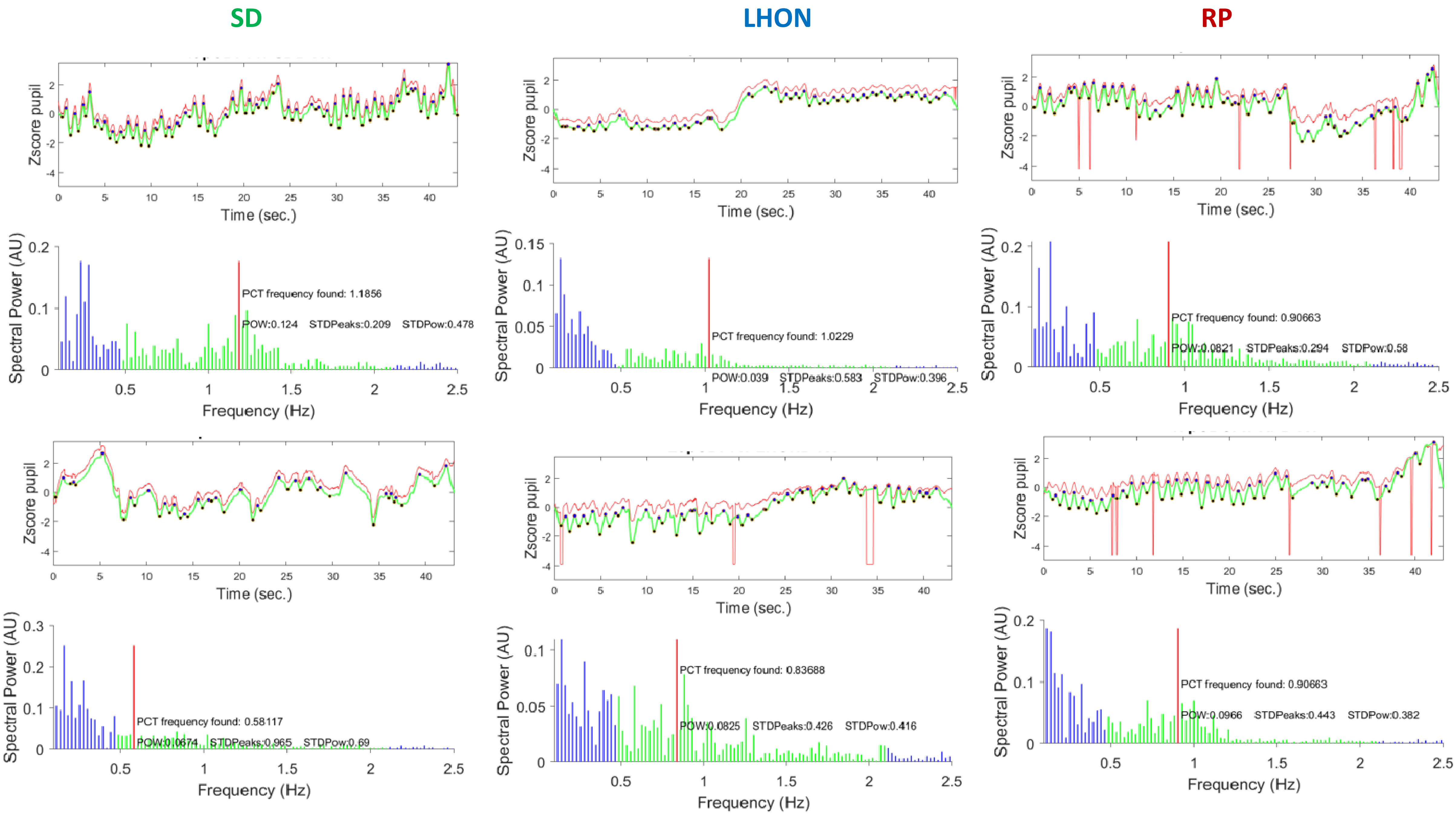

### Supplementary Table 1

| General | | Refraction RE | | | Refraction LE | | | Visual Acuity (LogMAR) | | | Pelli-Robson (logCS) | | | Summary | |
|---|---|---|---|---|---|---|---|---|---|---|---|---|---|---|---|
| Sex | Pathology | Sphere | CYL | Axis | Sphere | CYL | Axis | Right Eye | Left Eye | Binocular | Right Eye | Left Eye | Binocular | Age | Sex |
| M | HP | NA | NA | NA | NA | NA | NA | -0,3 | -0,2 | -0,3 | 1,65 | 1,65 | 1,65 | 37 +-10 | 8M / 6F |
| M | HP | NA | NA | NA | NA | NA | NA | -0,2 | -0,2 | -0,24 | 1,95 | 1,95 | 1,95 | Max 58 | |
| M | HP | -2,5 | 0 | 0 | -2,25 | -0,5 | 125 | 0 | -0,08 | -0,1 | 1,95 | 1,95 | 1,95 | Min 24 | |
| M | HP | 0 | 0 | 0 | 0 | 0 | 0 | -0,1 | -0,1 | -0,2 | 1,95 | 1,95 | 1,95 | | |
| F | HP | 0 | 0 | 0 | 0 | 0 | 0 | -0,02 | -0,1 | -0,12 | 1,95 | 1,95 | 1,95 | | |
| M | HP | -2 | -0,5 | 84 | -1,5 | -0,5 | 100 | -0,16 | -0,16 | -0,2 | 1,95 | 1,95 | 1,95 | | |
| F | HP | -10,25 | -1,25 | 157 | -9,5 | -1,75 | 172 | 0 | -0,04 | -0,1 | 1,95 | 1,95 | 1,95 | | |
| F | HP | 0 | 0 | 0 | 0 | 0 | 0 | -0,16 | -0,12 | -0,18 | 1,95 | 1,95 | 1,95 | | |
| F | HP | 0 | 0 | 0 | 0 | 0 | 0 | 0 | 0,04 | -0,04 | 1,95 | 1,95 | 1,95 | | |
| M | HP | 0 | 0 | 0 | 0 | 0 | 0 | -0,06 | -0,1 | -0,2 | 1,95 | 1,95 | 1,95 | | |
| M | HP | 0 | 0 | 0 | 0 | 0 | 0 | -0,16 | -0,2 | -0,26 | 1,95 | 1,95 | 1,95 | | |
| F | HP | -5,5 | -0,5 | 166 | -5 | -1,5 | 43 | 0 | -0,1 | -0,1 | 1,95 | 1,95 | 1,95 | | |
| F | HP | 0,5 | -0,25 | 106 | 0,75 | 0 | 0 | -0,2 | -0,12 | -0,26 | 1,95 | 1,95 | 1,95 | | |
| M | HP | 0 | 0 | 0 | 0 | 0 | 0 | -0,3 | -0,3 | -0,3 | 1,95 | 1,95 | 1,95 | | |
| M | SD | +0,5 | 0,25 | 80 | +0,5 | 0,25 | 175 | 0,86 | 0,76 | 0,78 | 1,65 | 1,65 | 1,65 | 38+-9 | 10M / 4F |
| M | SD | -0,25 | 0,25 | 65 | -0,75 | 0,25 | 5 | 0,3 | 0,4 | 0,2 | 1,5 | 1,35 | 1,65 | Max 55 | |
| F | SD | -2 | -0,5 | 75 | -2,25 | -1 | 90 | 1,02 | 1,02 | 1 | 1,2 | 1,2 | 1,2 | Min 21 | |
| F | SD | -1,25 | -0,5 | 80 | -1,75 | -0,75 | 25 | 0,86 | 0,74 | 0,74 | 1,5 | 1,65 | 1,5 | | |
| M | SD | -1,75 | -0,75 | 20 | -2 | -0,5 | 160 | 0,9 | 0,9 | 0,9 | 1,35 | 1,35 | 1,35 | | |
| M | SD | 1,75 | -1,5 | 25 | 2,5 | -2,25 | 170 | 1 | 1,1 | 1,1 | 1,35 | 1,35 | 1,35 | | |
| M | SD | -2,5 | -1,75 | 170 | -1,5 | -2 | 175 | 0,9 | 0,9 | 0,9 | 1,5 | 1,65 | 1,65 | | |
| M | SD | -1,5 | -1,75 | 180 | -1,25 | -3 | 180 | 0,94 | 0,96 | 0,86 | 1,5 | 1,2 | 1,5 | | |
| F | SD | 4 | -2 | 180 | 5 | -2,25 | 5 | 1,04 | 1,18 | 1 | 1,5 | 0,9 | 1,35 | | |
| M | SD | 5,5 | -1,75 | 15 | 7 | -0,5 | 7 | 1,26 | 1,16 | - | 0,75 | 0,6 | DM | | |
| M | SD | 0 | -1,5 | 170 | -1,5 | -0,5 | 170 | 1,04 | 0,92 | 0,92 | 1,05 | 1,2 | 1,35 | | |
| M | SD | -3 | -0,75 | 105 | -3,5 | -0,25 | 140 | 1 | 1,16 | 1,02 | 1,35 | 1,2 | 1,35 | | |
| F | SD | 0,5 | -1,25 | 25 | 0,75 | -1,25 | 150 | 1 | 1 | 1 | 1,05 | 1,05 | 1,2 | | |
| M | SD | 0,75 | -0,75 | 90 | 0,75 | -0,5 | 115 | 1,1 | 1,3 | 1,2 | 1,2 | 1,35 | 1,35 | | |
| F | RP | -1,75 | 2 | 5 | -1,75 | 2,5 | 165 | 0,26 | 0,34 | 0,22 | 0,9 | 0,9 | 1,2 | 41+-10 | 8M / 6 F |
| M | RP | -7,25 | -2,25 | 20 | -8 | -2 | 175 | 0,38 | 0,42 | 0,42 | 1,65 | 1,35 | 1,65 | Max 58 | 11 |
| M | RP | -4,75 | 3,25 | 130 | -6,25 | 2,5 | 25 | 0,72 | 0,76 | 0,66 | 0,45 | 0,15 | 0,45 | Min 24 | |
| M | RP | -5,5 | -0,25 | 160 | -4 | -1,25 | 20 | 0,04 | 0,04 | 0,02 | 1,65 | 1,65 | 1,65 | | |
| M | RP | 0 | 0,5 | 100 | -0,5 | 0,5 | 110 | -0,04 | -0,08 | -0,08 | 1,65 | 1,65 | 1,65 | | |
| M | RP | -6 | 1,5 | 10 | -3,5 | 0,75 | 180 | 0,84 | 1,02 | 0,78 | 0,45 | 0,3 | 0,45 | | |
| M | RP | -0,5 | 1 | 125 | -2 | 1,25 | 10 | 0,72 | 0,7 | 0,68 | 0,45 | 0,6 | 0,6 | | |
| F | RP | -2,75 | -1,5 | 85 | -3,25 | -1,5 | 80 | 0,06 | 0,22 | 0 | 1,65 | 1,65 | 1,65 | | |
| M | RP | -1,25 | -1,75 | 90 | 0,75 | -0,5 | 115 | 0,28 | -0,04 | -0,04 | 1,65 | 1,65 | 1,65 | | |
| F | RP | -0,75 | -1,75 | 75 | -0,75 | -2 | 105 | 0,34 | 0,22 | 0,3 | 1,65 | 1,5 | 1,65 | | |
| F | RP | -6 | -2,5 | 20 | -6,5 | -2,25 | 155 | 0,34 | 0,44 | 0,34 | 1,35 | 1,05 | 1,35 | | |
| F | RP | -6 | -2,25 | 15 | -6 | -1 | 135 | 0,22 | 0,12 | 0,1 | 1,65 | 1,5 | 1,65 | | |
| M | RP | -3,5 | -1 | 0 | -2,25 | -1,25 | 5 | 0,36 | 0,12 | 0,1 | 1,5 | 1,65 | 1,65 | | |
| F | RP | NA | NA | NA | NA | NA | NA | 0,3 | 0,4 | 0,3 | 1,65 | 1,65 | 1,65 | | |
| M | LHON | -0,5 | 0,75 | 80 | -0,25 | 1,75 | 90 | 0 | 0,1 | 0 | 1,05 | 1,05 | 1,05 | 33 +-7 | 5M / 4F |
| F | LHON | -1,5 | -0,25 | 15 | -1 | -0,25 | 15 | 0,32 | - | 0,32 | 0,45 | DM | 0,45 | Max 42 | |
| M | LHON | 1,25 | NA | NA | 1,25 | NA | NA | 1,52 | 1,48 | 1,48 | 0 | 0,45 | 0,45 | Min 20 | |
| F | LHON | 2,5 | 0 | 0 | 3,25 | -0,5 | 135 | 1,4 | 1,3 | 1,4 | 0,75 | 1,05 | 0,75 | | |
| F | LHON | -0,75 | -0,25 | 130 | -0,75 | -0,5 | 15 | 1,02 | 1,1 | 0,92 | 1,05 | 1,05 | 1,35 | | |
| M | LHON | -3 | -1,25 | 5 | -2,75 | -0,25 | 125 | 1,44 | 1,32 | 1,36 | 0,6 | 0,45 | 0,75 | | |
| M | LHON | 0,25 | 0 | 0 | 0 | 0 | 0 | 1,04 | 0,24 | 0,44 | 0,15 | 0,15 | 0,15 | | |
| F | LHON | -0,5 | -0,75 | 100 | -1 | -0,5 | 75 | 1,5 | 1,26 | 1,32 | 0,15 | 0,6 | 0,45 | | |
| M | LHON | 0 | -0,75 | 10 | 0,25 | -1 | 10 | 1 | 0,96 | 0,96 | 1,35 | 1,5 | 1,65 | | |

Summary statistics of the participants

HP : Healthy participants

SD : Stargardt disease

RP: Retinitis Pigmentosa

LHON : Leber Heriditary Optic Neuropathy

### Supplementary table 2: Statistical results

ANOVA Factors: PATHO {HP, RP, SD, LHON} STI {FULL, RING, DISK } COL {Blue, Red, Green, Grey}

Variables used for Analyzes of Variance:
'Power' 'Frequency' 'Var Freq' 'MeanFFT' 'FFTavg' 'StdFFT' 'Period' 'MeanAmp' 'Var Amp' 'MoyPfft' 'BLK Ratio' 'Slope'

#### ANOVA summary

| **Source Power** | Sum Sq. | d.f. | Mean Sq. | F | Prob>F |
|---|---|---|---|---|---|
| PATHO | 1,167 | 3 | 0,39 | 37,57 | 0,00000 |
| Sti | 0,437 | 2 | 0,22 | 21,12 | 0,00000 |
| Col | 0,269 | 3 | 0,09 | 8,66 | 0,00001 |
| PATHO*Sti | 0,101 | 6 | 0,02 | 1,62 | 0,13900 |
| PATHO*Col | 0,107 | 9 | 0,01 | 1,14 | 0,33093 |
| Sti*Col | 0,011 | 6 | 0,00 | 0,17 | 0,98380 |
| PATHO*Sti*Col | 0,054 | 18 | 0,00 | 0,29 | 0,99828 |

| **Source Frequency** | Sum Sq. | d.f. | Mean Sq. | F | Prob>F |
|---|---|---|---|---|---|
| PATHO | 2,417 | 3 | 0,81 | 29,34 | 0,00000 |
| Sti | 0,132 | 2 | 0,07 | 2,40 | 0,09201 |
| Col | 1,362 | 3 | 0,45 | 16,53 | 0,00000 |
| PATHO*Sti | 0,610 | 6 | 0,10 | 3,70 | 0,00134 |
| PATHO*Col | 0,544 | 9 | 0,06 | 2,20 | 0,02119 |
| Sti*Col | 0,381 | 6 | 0,06 | 2,31 | 0,03289 |
| PATHO*Sti*Col | 0,308 | 18 | 0,02 | 0,62 | 0,88234 |

| **Source Var Freq** | Sum Sq. | d.f. | Mean Sq. | F | Prob>F |
|---|---|---|---|---|---|
| PATHO | 4,162 | 3 | 1,39 | 30,45 | 0,00000 |
| Sti | 1,329 | 2 | 0,66 | 14,59 | 0,00000 |
| Col | 1,047 | 3 | 0,35 | 7,66 | 0,00005 |
| PATHO*Sti | 0,527 | 6 | 0,09 | 1,93 | 0,07480 |
| PATHO*Col | 1,128 | 9 | 0,13 | 2,75 | 0,00391 |
| Sti*Col | 0,017 | 6 | 0,00 | 0,06 | 0,99897 |
| PATHO*Sti*Col | 0,205 | 18 | 0,01 | 0,25 | 0,99940 |

| **Source MeanFFT** | Sum Sq. | d.f. | Mean Sq. | F | Prob>F |
|---|---|---|---|---|---|
| PATHO | 0,006 | 3 | 0,00 | 20,15 | 0,00000 |
| Sti | 0,002 | 2 | 0,00 | 9,55 | 0,00009 |
| Col | 0,003 | 3 | 0,00 | 10,55 | 0,00000 |
| PATHO*Sti | 0,001 | 6 | 0,00 | 1,74 | 0,10951 |
| PATHO*Col | 0,001 | 9 | 0,00 | 1,10 | 0,36024 |
| Sti*Col | 0,000 | 6 | 0,00 | 0,33 | 0,92018 |
| PATHO*Sti*Col | 0,000 | 18 | 0,00 | 0,25 | 0,99933 |

| **Source FFTavg** | Sum Sq. | d.f. | Mean Sq. | F | Prob>F |
|---|---|---|---|---|---|
| PATHO | 0,000 | 3 | 0,00 | 6,80 | 0,00017 |
| Sti | 0,000 | 2 | 0,00 | 2,11 | 0,12223 |
| Col | 0,000 | 3 | 0,00 | 7,06 | 0,00012 |
| PATHO*Sti | 0,000 | 6 | 0,00 | 1,80 | 0,09819 |
| PATHO*Col | 0,000 | 9 | 0,00 | 1,37 | 0,19862 |
| Sti*Col | 0,000 | 6 | 0,00 | 0,21 | 0,97230 |
| PATHO*Sti*Col | 0,000 | 18 | 0,00 | 0,54 | 0,94034 |

| **Source StdFFT** | Sum Sq. | d.f. | Mean Sq. | F | Prob>F |
|---|---|---|---|---|---|
| PATHO | 0,023 | 3 | 0,01 | 35,33 | 0,00000 |
| Sti | 0,009 | 2 | 0,00 | 21,13 | 0,00000 |
| Col | 0,006 | 3 | 0,00 | 9,52 | 0,00000 |
| PATHO*Sti | 0,002 | 6 | 0,00 | 1,80 | 0,09786 |
| PATHO*Col | 0,002 | 9 | 0,00 | 1,01 | 0,43150 |
| Sti*Col | 0,000 | 6 | 0,00 | 0,31 | 0,93355 |
| PATHO*Sti*Col | 0,001 | 18 | 0,00 | 0,23 | 0,99963 |

| **Source Period** | Sum Sq. | d.f. | Mean Sq. | F | Prob>F |
|---|---|---|---|---|---|
| PATHO | 2,563 | 3 | 0,85 | 27,41 | 0,00000 |
| Sti | 0,261 | 2 | 0,13 | 4,19 | 0,01585 |
| Col | 0,794 | 3 | 0,26 | 8,49 | 0,00002 |
| PATHO*Sti | 1,051 | 6 | 0,18 | 5,62 | 0,00001 |
| PATHO*Col | 0,873 | 9 | 0,10 | 3,11 | 0,00123 |
| Sti*Col | 0,069 | 6 | 0,01 | 0,37 | 0,89945 |
| PATHO*Sti*Col | 0,246 | 18 | 0,01 | 0,44 | 0,97909 |

| **Source Mean Amp** | Sum Sq. | d.f. | Mean Sq. | F | Prob>F |
|---|---|---|---|---|---|
| PATHO | 28,085 | 3 | 9,36 | 33,59 | 0,00000 |
| Sti | 9,910 | 2 | 4,96 | 17,78 | 0,00000 |
| Col | 9,031 | 3 | 3,01 | 10,80 | 0,00000 |
| PATHO*Sti | 2,789 | 6 | 0,46 | 1,67 | 0,12738 |
| PATHO*Col | 2,194 | 9 | 0,24 | 0,87 | 0,54805 |
| Sti*Col | 0,735 | 6 | 0,12 | 0,44 | 0,85224 |
| PATHO*Sti*Col | 1,045 | 18 | 0,06 | 0,21 | 0,99984 |

| **Source Var Amp** | Sum Sq. | d.f. | Mean Sq. | F | Prob>F |
|---|---|---|---|---|---|
| PATHO | 0,380 | 3 | 0,13 | 4,30 | 0,00529 |
| Sti | 0,028 | 2 | 0,01 | 0,47 | 0,62610 |
| Col | 0,152 | 3 | 0,05 | 1,72 | 0,16240 |
| PATHO*Sti | 0,068 | 6 | 0,01 | 0,39 | 0,88748 |
| PATHO*Col | 0,928 | 9 | 0,10 | 3,50 | 0,00035 |
| Sti*Col | 0,211 | 6 | 0,04 | 1,19 | 0,31066 |
| PATHO*Sti*Col | 0,671 | 18 | 0,04 | 1,26 | 0,20816 |

| **Source MoyPfft** | Sum Sq. | d.f. | Mean Sq. | F | Prob>F |
|---|---|---|---|---|---|
| PATHO | 6609374,2 | 3 | 2203124,75 | 0,33 | 0,80113 |
| Sti | 3049770,6 | 2 | 1524885,31 | 0,23 | 0,79397 |
| Col | 1105853,1 | 3 | 368617,70 | 0,06 | 0,98265 |
| PATHO*Sti | 3129074,6 | 6 | 521512,43 | 0,08 | 0,99812 |
| PATHO*Col | 34846394,8 | 9 | 3871821,64 | 0,59 | 0,80873 |
| Sti*Col | 4946805,1 | 6 | 824467,52 | 0,12 | 0,99331 |
| PATHO*Sti*Col | 25625788,2 | 18 | 1423654,90 | 0,22 | 0,99979 |

| **Source BLK Ratio** | Sum Sq. | d.f. | Mean Sq. | F | Prob>F |
|---|---|---|---|---|---|
| PATHO | 0,120 | 3 | 0,04 | 2,10 | 0,09918 |
| Sti | 0,018 | 2 | 0,01 | 0,48 | 0,61721 |
| Col | 0,032 | 3 | 0,01 | 0,56 | 0,63892 |
| PATHO*Sti | 0,025 | 6 | 0,00 | 0,22 | 0,97140 |
| PATHO*Col | 0,144 | 9 | 0,02 | 0,85 | 0,57477 |
| Sti*Col | 0,066 | 6 | 0,01 | 0,58 | 0,74641 |
| PATHO*Sti*Col | 0,082 | 18 | 0,00 | 0,24 | 0,99954 |

| **Source Slope** | Sum Sq. | d.f. | Mean Sq. | F | Prob>F |
|---|---|---|---|---|---|
| PATHO | 0,021 | 3 | 0,01 | 7,43 | 0,00007 |
| Sti | 0,001 | 2 | 0,00 | 0,54 | 0,58291 |
| Col | 0,010 | 3 | 0,00 | 3,45 | 0,01658 |
| PATHO*Sti | 0,003 | 6 | 0,00 | 0,56 | 0,76213 |
| PATHO*Col | 0,009 | 9 | 0,00 | 1,10 | 0,36405 |
| Sti*Col | 0,007 | 6 | 0,00 | 1,23 | 0,29077 |
| PATHO*Sti*Col | 0,014 | 18 | 0,00 | 0,85 | 0,63764 |
